# Domiciliary high-flow nasal cannula oxygen therapy for stable hypercapnic COPD patients: a prospective, multicenter, open-label, randomized controlled trial

**DOI:** 10.1101/2021.05.21.21257508

**Authors:** Kazuma Nagata, Takeo Horie, Naohiko Chohnabayashi, Torahiko Jinta, Ryosuke Tsugitomi, Akira Shiraki, Fumiaki Tokioka, Toru Kadowaki, Akira Watanabe, Motonari Fukui, Takamasa Kitajima, Susumu Sato, Toru Tsuda, Nobuhito Kishimoto, Hideo Kita, Yoshihiro Mori, Masayuki Nakayama, Tomomasa Tsuboi, Makoto Yoshida, Osamu Hataji, Satoshi Fuke, Michiko Kagajo, Hiroki Nishine, Hiroyasu Kobayashi, Hiroyuki Nakamura, Miyuki Okuda, Sayaka Tachibana, Shohei Takata, Hisayuki Osoreda, Kenichi Minami, Takashi Nishimura, Tadashi Ishida, Jiro Terada, Naoko Takeuchi, Yasuo Kohashi, Hiromasa Inoue, Yoko Nakagawa, Takashi Kikuchi, Keisuke Tomii, FLOCOP study investigators

**Affiliations:** Department of Respiratory Medicine, Kobe City Medical Center General Hospital, Kobe, Hyogo, Japan.; Department of Respiratory Medicine, Japanese Red Cross Maebashi Hospital, Maebashi, Gunma, Japan.; Division of Pulmonary Medicine, Thoracic Center, St. Luke’s International Hospital, Chuo-ku, Tokyo, Japan.; Department of Respiratory Medicine, Ogaki Municipal Hospital, Ogaki, Gifu, Japan.; Department of Respiratory Medicine, Kurashiki Central Hospital, Kurashiki, Okayama, Japan.; Department of Pulmonary Medicine, National Hospital Organization Matsue Medical Center, Matsue, Shimane, Japan.; Department of Respiratory Medicine, National Hospital Organization Ehime Medical Center, Toon, Ehime, Japan.; Respiratory Disease Center, Tazuke Kofukai Foundation, Medical Research Institute, Kitano Hospital, Osaka, Osaka, Japan; Department of Respiratory Medicine, Graduate School of Medicine, Kyoto University, Kyoto, Kyoto, Japan; Kirigaoka Tsuda Hospital, Kitakyushu, Fukuoka, Japan.; Department of Respiratory Medicine, Takamastu Municipal Hospital, Takamatsu, Kagawa, Japan.; Department of Respiratory Medicine, Takatsuki Red-Cross Hospital, Takatsuki, Osaka, Japan; Department of Respiratory Medicine, KKR Takamatsu Hospital, Takamatsu, Kagawa, Japan; Division of Pulmonary Medicine, Department of Medicine, Jichi Medical University, Shimotsuke, Tochigi, Japan.; Department of Respiratory Medicine, National Hospital Organization Minami Kyoto Hospital, Joyo, Kyoto, Japan; Department of Respiratory Medicine, National Hospital Organization Fukuoka National Hospital, Fukuoka, Fukuoka, Japan.; Respiratory Center, Matsusaka municipal hospital, Matsusaka, Mie, Japan.; Department of Respiratory Medicine, KKR Sapporo Medical Center, Sapporo, Hokkaido, Japan.; Division of Respiratory Medicine, Department of Internal Medicine, St. Marianna University School of Medicine, Kawasaki, Kanagawa, Japan.; Respiratory Center, Suzuka General Hospital, Suzuka, Mie, Japan.; Department of Respiratory Medicine, Sakaide City Hospital, Sakaide, Kagawa, Japan.; Osaka Anti-tuberculosis Association Osaka Hospital, Neyagawa, Osaka, Japan.; Department of Respiratory Medicine, Ehime Prefectural Central Hospital, Matsuyama, Ehime, Japan.; Department of Respiratory Medicine, National Hospital Organization Fukuokahigashi Medical Center, Koga, Fukuoka, Japan.; Department of Respiratory Medicine, NHO Yamaguchi-Ube Medical Center, Ube, Yamaguchi, Japan.; Department of Respiratory Medicine, Ishikiriseiki Hospital, Higashi-Osaka, Osaka, Japan.; Department of Respiratory Medicine, Kyoto Katsura Hospital, Kyoto, Kyoto, Japan.; Department of Respirology, Graduate School of Medicine, Chiba University, Chiba, Chiba, Japan.; Department of Internal Medicine, National Hospital Organization Kinki-Chuo Chest Medical Center, Sakai, Osaka, Japan.; Department of Respiratory Medicine, HARUHI Respiratory Medical Hospital, Kiyosu, Aichi, Japan.; Department of Pulmonary Medicine, Graduate School of Medical and Dental Sciences, Kagoshima University, Kagoshima, Kagoshima, Japan.; Division of Medical Statistics, Translational Research Center for Medical Innovation, Foundation for Biomedical Research and Innovation at Kobe, Kobe, Hyogo, Japan.

**Keywords:** high-flow nasal cannula, chronic hypercapnic respiratory failure, chronic obstructive pulmonary disease, long-term oxygen therapy

## Abstract

**Background:** The effectiveness of the domiciliary use of high-flow nasal cannula oxygen therapy (HFNC) in patients with chronic hypercapnic respiratory failure due to chronic obstructive pulmonary disease (COPD) remains controversial.

**Objectives:** To investigate the efficacy and safety of domiciliary HFNC use in stable hypercapnic COPD patients.

**Methods:** This multicenter, open-label, randomized controlled trial enrolled patients with stable hypercapnic COPD. Over 52 weeks, we compared long-term oxygen therapy (LTOT) alone versus domiciliary HFNC plus LTOT (HFNC/LTOT). The primary endpoint was the frequency of moderate/severe COPD exacerbations. We also compared changes from baseline levels in arterial blood gas, SpO_2_, pulmonary function, health-related quality of life (HRQOL), and a six-minute walk test.

**Results:** We enrolled 104 patients in total; from these, we removed mismatching patients and then assigned 49 and 50 patients to HFNC/LTOT and LTOT groups, respectively, for safety analysis; 47 and 46 patients for HFNC/LTOT and LTOT groups, respectively, for efficacy analysis. Thirty-seven (79%) and 41 patients (89%) in HFNC/LTOT and LTOT, respectively, completed the final evaluation. HFNC significantly reduced the frequency of COPD exacerbations and prolonged the duration without moderate or severe COPD exacerbations over the 52-week study period (*p* = 0.002, *p* = 0.032, respectively). The adjusted odds ratios (95% confidence intervals [CIs]) of the frequency of COPD exacerbations in LTOT against HFNC/LTOT was 2.85 (1.48, 5.47). The median survival time (95% CI) to the first COPD exacerbation with moderate or severe for the LTOT group was 25 weeks (14.1, 47.4); however, the HFNC/LTOT group did not reach the median survival time. HFNC also caused statistically significant differences (*p* < 0.05) in the SpO_2_, FVC (%FVC), and FEV1 (%FEV1); however, these improvements were transient. There were no other improvements in arterial blood gas, pulmonary function, HRQOL, or six-minute walk test parameters. In addition, no safety concerns were identified for HFNC.

**Conclusions:** HFNC may be a reasonable therapeutic choice in stable hypercapnic COPD patients with a history of exacerbations.

*Trial registration number*: UMIN000028581, NCT03282019 (http://www.umin/ac.jp, https://clinicaltrials.gov/)

## INTRODUCTION

Advanced-stage chronic obstructive pulmonary disease (COPD) is characterized by severe airflow limitations, severely limited performance, and systemic complications.[1] This disease progressively leads to chronic respiratory insufficiency, often characterized by hypercapnia or hypoxia. Once patients develop chronic hypercapnic respiratory failure, their prognosis worsens, and they experience a deterioration in their condition.[2] Moreover, they are more likely to develop exacerbations, with poor outcomes in terms of mortality and recurrence of exacerbations.[3]

One treatment option for patients with chronic hypercapnic respiratory failure is noninvasive ventilation (NIV) in an at-home setting. Previous randomized controlled trials have demonstrated that NIV use improved physiological parameters and survival rates and reduces readmission rates following hospitalization for COPD exacerbations.[4–6] However, certain barriers to NIV use can compromise treatment compliance and even lead to its failure, including interface discomfort, sleep disturbances, and intolerability caused by patient-ventilator asynchrony.[7–9] Therefore, there is an unmet need for alternative treatment strategies that are both well tolerated and easy to administer.

High-flow nasal cannula oxygen therapy (HFNC) is a gas delivery system providing heated, humidified air by nasal cannula, with supplemental oxygen as required. HFNC creates a lower level of positive airway pressure than NIV does, has a washout effect in pharyngeal dead space, decreases inspiratory resistance, and can improve mucus clearance.[10–13] HFNC has been widely studied in adult intensive care units for treating patients with acute hypoxemic respiratory failure[14, 15] and after extubation.[16, 17] In addition, there is growing evidence that HFNC can be beneficial in patients with chronic respiratory failure.

In a recent meta-analysis of several randomized trials, HFNC was shown to reduce the arterial partial pressure of carbon dioxide (PaCO2) and the number of exacerbations in stable COPD patients and to increase health-related quality of life (HRQOL).[18] However, most current studies have focused on the short-term effects of HFNC, with sparse data on its long-term effects. In a randomized controlled trial assessing 12 months of HFNC use in an at-home setting in Denmark, consistent use of HFNC reduced the frequency of COPD exacerbations and hospitalizations.[19] However, this Danish study recruited COPD patients with chronic hypoxemic respiratory failure, with only half of them being hypercapnic at inclusion. In another randomized trial, HFNC significantly reduced the number of exacerbation days and increased the time to first exacerbation in a mixed population of COPD and bronchiectasis patients for 12 months. However, it is still unclear whether long-term use of HFNC can be effective and safe for the most severe COPD patients with chronic hypercapnic respiratory failure.

In the current study, it was hypothesized that the long-term use of HFNC in COPD patients with chronic hypercapnic respiratory failure would reduce the number of COPD exacerbations and improve mortality rates, HRQOL, and physiological parameters.

## MATERIALS and METHODS

This trial was an investigator-initiated, multicenter, open-label, parallel-group randomized controlled clinical trial with a 1:1 allocation to either long-term oxygen therapy (LTOT) alone or domiciliary HFNC plus LTOT (HFNC/LTOT). Patients were recruited from 42 hospitals in Japan. The inclusion and exclusion criteria are presented in Table E1. Briefly, men and women aged 40 years or older with daytime hypercapnia (PaCO2 ≥45 mmHg and pH ≥7.35) and Global Initiative for Obstructive Lung Disease (GOLD)[20] stage 2-4 disease receiving LTOT for at least 16 hours per day for at least 1 month prior to providing informed consent were recruited. Patients were required to have had an exacerbation (moderate or severe; judged by the investigators) within the past 1 year. Patients were in stable condition, free from a COPD exacerbation (of any severity) within the 4 weeks prior to enrollment. Patients who had used nocturnal NIV within the past 4 weeks or HFNC within the past 1 year were excluded. Patients who had any history of obstructive sleep apnea syndrome (OSAS) or were highly suspected to have OSAS based on clinical findings were also excluded. All patients were receiving optimal medical therapy according to GOLD guidelines[20] in addition to LTOT at the time of enrollment. Changes to medication, commencement of rehabilitation, or use of NIV were not permitted throughout the study period, except temporarily (less than 14 days) in patients with COPD exacerbations. This trial was conducted in accordance with the principles of the Declaration of Helsinki and the Ethical Guidelines for Medical and Health Research Involving Human Subjects (Japanese Ministry of Health, Labour and Welfare). The study protocol was approved by the Kobe University Clinical Research Ethical Committee (C180079), and written informed consent was obtained from all patients before participating in the trial. This study is registered at ClinicalTrials.gov (NCT03282019) and the University Hospital Medical Information Network Clinical Trials Registry (UMIN-CTR; UMIN000028581). This trial was registered on the UMIN website prior to enrollment of the first study participant.

### Interventions

Patients who met the eligibility criteria were randomized to receive either LTOT alone or domiciliary HFNC/LTOT. Randomization was performed using a permuted block method at the Translational Research Informatics Center in Kobe, Japan. All study participants were instructed to continue their prescribed LTOT during the daytime. Study participants allocated to the HFNC/LTOT group were instructed to use HFNC for at least 4 hours per night during sleep at flow rates of 30–40 L/min in addition to LTOT. HFNC was administered using the myAIRVO2® device, which provides humidification and high-flow medical gas via an Optiflow nasal cannula interface (Fisher & Paykel Healthcare, Auckland, New Zealand). The investigator was permitted to adjust the nocturnal oxygen flow rate in order to maintain a peripheral oxygen saturation (SpO2) of ≥88%. If subjects reported discomfort with HFNC use, the investigator could down-titrate the flow rate to a minimum of 20 L/min. Training was provided before the start of HFNC by trained doctors and nurses in order to habituate study participants to HFNC. Study participants were also allocated time to adapt themselves to the interface and machine and were trained to operate and maintain the machine by themselves at home. Study participants were discharged after they were comfortable with using the machine and all at-home study procedures.

### Follow-up and Measurements

All participants were followed throughout the 52-week study period, and regular study visits were scheduled every 4 weeks. Participants were instructed to complete a daily diary, and records were reviewed at each study visit to determine whether respiratory symptoms met the criteria for COPD exacerbation. At every visit, doctors also verified the COPD treatment regimen and adherence with HFNC (only for the HFNC/LTOT group) and evaluated for any adverse events (AEs), hospital admissions, and SpO2. In addition, participants were assessed for HRQOL, sleep quality, and dyspnea and underwent pulmonary function testing (PFT) and the 6-minute walk test (6MWT) at 12, 24, and 52 weeks and at admission, if needed. A deviation of up to 1 week was permitted.

Participants completed a daily diary to record any increases in upper respiratory tract symptoms (e.g., nasal discharge, sore throat), increases in lower respiratory tract symptoms (e.g., dyspnea, sputum, cough, wheezing), fever, or use of systemic corticosteroids or antibiotics.[21] A COPD exacerbation was diagnosed if one of the following symptom patterns were experienced for at least 2 consecutive days: either two or more of three major symptoms (increases in dyspnea, sputum purulence, or sputum volume) or any one major symptom in addition to one of the following minor symptoms: increase in nasal discharge, wheezing, sore throat, cough, or fever.[22] A mild COPD exacerbation was defined as being resolved without use of systemic corticosteroids or antibiotics. A moderate COPD exacerbation was defined as necessitating treatment with systemic corticosteroids and/or antibiotics. A severe COPD exacerbation was defined as requiring hospitalization, including an emergency admission.

Adherence was confirmed by the myAIRVO2® usage time obtained directly from the log record of the device. HRQOL was assessed using the St. George’s Respiratory Questionnaire for COPD (SGRQ-C)[23] and the Severe Respiratory Insufficiency Questionnaire (SRI) as disease specific questionnaires.[24] Quality-adjusted life years (QALYs) were evaluated using a five-level version of the EuroQol five-dimensional questionnaire (EQ-5D-5L)[25] to assess the economic benefits of HFNC. Sleep quality was assessed using the Japanese version of the Pittsburgh Sleep Quality Index (PSQI-J).[26, 27] The subjective severity of dyspnea was measured using the modified Medical Research Council (mMRC) scale.[28] PFTs were performed by trained operators in accordance with the international guidelines.[29] Predicted pulmonary function test values were calculated based on Japanese Respiratory Society guidelines.[30]

Arterial blood samples and SpO2 values measured using pulse oximetry were assessed in the morning with patients in the supine position for 5 minutes under oxygen therapy at the flow rate prescribed by their treating physician. The 6MWT was performed while receiving oxygen, according to the ATS guidelines.[31]

### Study Outcomes

We defined the primary and secondary study endpoints as follows:

1. Primary endpoint Counts per 52 weeks or frequency of moderate or severe COPD exacerbations
2. Secondary endpoints

1. Time to first moderate/severe COPD exacerbation
2. Time to death by all causes: overall survival (OS)
3. Frequency per 52 weeks of COPD exacerbations with all severities (mild/moderate/severe) or severe severity only 1.
4. Changes in the total SGRQ-C score and each component score (i.e., symptom, activity, and impact scores)
5. QALY calculated by the EQ-5D-5L score
6. Changes in HRQOL scores (SRI, PSQI-J, mMRC scale)
7. Changes in arterial blood gas measurements (pH, PaCO2, PaO2, HCO3-, BE)
8. Changes in SpO2
9. Changes in pulmonary functions (VC, %VC, FVC, %FVC, FEV1, %FEV1, FEV1/FVC, FEV1%, DLCO, %DLCO)
10. Changes in 6MWT measurements (walk distance, SpO2 at pre and post-test, modified Borg scale at post-test)
11. Time to commencement of long-term NIV
12. AEs caused by myAIRVO2® therapy

In these definitions, changes represented the differences in values between baseline and at each scheduled measurement.

### Sample Size

The required sample size was determined to be 120 patients in total, including a nearly 10% rate of potential dropouts. We determined this sample size based on patient recruitment feasibility and with consideration that the evaluable size per treatment should be 53 patients at a minimum. We determined that this size could detect an effect size of 1.0, which was the difference in the mean count of COPD exacerbations per year (moderate or severe) between the HFNC/LTOT and LTOT groups, with a 90% power, a two-tailed type I error rate of 5%, and an estimated population standard deviation of 1.58.

### Statistical Analysis

We expected that many patients would not experience a COPD exacerbation during the study, leading to a data distribution with many zero counts and a very skewed distribution. To remedy this issue, we used a generalized linear regression model (GLRM) in which the distribution family was a negative binomial distribution, with treatment, gender, and GOLD stage as factors and age as a covariate. The model’s base level was HFNC/LTOT, male, and GOLD stage 2. We also carried out a goodness-of-fit test (chi-squared test) for fitting data.

We used Kaplan-Meier curve analysis for the secondary endpoints of 1) time to first moderate/severe COPD exacerbation and 2) OS, and examined a null hypothesis that the two treatments curves would be identical using log-rank testing. We also calculated median survival times with 95% confidence intervals (CIs) if the curves reached the median survival time.

To compare the adjusted mean or the least-squared mean (LSM) of the effects of HFNC/LTOT and LTOT at each measurement week, we used a Mixed Models Repeated Measurements (MMRM) technique,[12] in which the fixed effects were treatment, time, and interaction term and the random effect was the patient, and we assumed an unstructured variance-covariance matrix for time. We applied the MMRM technique for secondary endpoints 4); 6), except the mMRC scale; and 8).

Using *t*-tests, we compared changes in secondary endpoints 7), 9), and 10) between the treatment groups. If the *p*-value was less than 0.05 for any item in 7), we confirmed the result using the MMRM technique and considered that the MMRM’s result should give a conclusion for the item. We also used the Fisher’s exact test for categorical data from the mMRC scale.

To evaluate the QALYs calculated by the EQ-5D-5L, we used a covariance (ANCOVA) analysis with a factor of treatments and a covariate of baseline scores.

These analyses were performed using SAS 9.4 and Ri 386 3.4.3. We set the significance level at 0.05 for statistical tests and did not adjust the significance level for repeated tests because this study had an explanatory purpose. We defined the safety set (SS) and full analysis set (FAS) as the data sets intended to analyze “patient backgrounds and AE” and “primary and secondary endpoints,” respectively.

## RESULTS

### Patients

We initially enrolled 104 patients but excluded 5 due to a lack of study treatments, resulting in 99 patients in the SS. Because six patients did not provide self-records for COPD exacerbations, we removed them from the Safety analysis data Set (SS) so that there were 93 patients in the Full Analysis data Set (FAS) for efficacy analysis. All patients in the SS and FAS provided safety and efficacy data for our intention-to-treat basis analysis.

The number of the HFNC/LTOT and LTOT groups in the FAS were 47 and 46. At the 52nd week, 37 out of 47 and 41 out of 46 patients remained for the final evaluation.

Table 1 shows patient demographic data, smoking history, GOLD stages, and currently prescribed drugs, with no statistically significant differences (SSDs) in these variables between the two treatment groups.

**Table 1.** Patient demographic data, smoking status, GOLD stage, and prescribed drugs. HFNC, high-flow nasal cannula oxygen therapy; LTOT, long-term oxygen therapy; BMI, body mass index; GOLD, Global Initiative for Obstructive Lung Disease; LAMA, long-acting muscarinic agent; LABA, long-acting beta agonist; SD, standard deviation

|  |  |  | HFNC/LTOT | LTOT |  |
| --- | --- | --- | --- | --- | --- |
| Characteristic |  |  | n (%) | n (%) | P-value |
| Sex | Male |  | 44 (89.8) | 44 (88.0) | 1.000 |
|  | Female |  | 5 (10.2) | 6 (12.0) | . |
| Age | Mean |  | 72.9 | 75.16 | 0.114 |
|  | SD |  | 7.43 | 6.67 | . |
| BMI | Mean |  | 20.21 | 20.38 | 0.815 |
|  | SD |  | 3.57 | 3.64 | . |
| Smoking history | Current |  | 0 (0.0) | 2 (4.0) | 0.495 |
|  | Past |  | 48 (98.0) | 48 (96.0) | . |
|  | Never |  | 1 (2.0) | 0 (0.0) | . |
|  | Number of cigarettes per day | Mean | 31.67 | 30 | 0.565 |
|  |  | SD | 14.7 | 13.85 | . |
|  | Years | Mean | 39.63 | 39.84 | 0.918 |
|  |  | SD | 11.17 | 9.35 | . |
| GOLD stage | 2 |  | 1 (2.0) | 4 (8.0) | 0.445 |
|  | 3 |  | 10 (20.4) | 11 (22.0) | . |
|  | 4 |  | 38 (77.6) | 35 (70.0) | . |
| LAMA § | N |  | 1 (2.0) | 7 (14.0) | 0.0594 |
|  | Y |  | 48 (98.0) | 43 (86.0) | . |
| LABA § | N |  | 2 (4.1) | 4 (8.0) | 0.678 |
|  | Y |  | 47 (95.9) | 46 (92.0) | . |
| Inhaled steroids § | N |  | 22 (44.9) | 19 (38.0) | 0.544 |
|  | Y |  | 27 (55.1) | 31 (62.0) | . |

§ : Duplicate counted

### Treatments

The mean (SD: standard deviation) for the use of HFNC/LTOT was 7.3 (3.0) hours per day. During the study, including baseline, the means (SD) of oxygen flow rate (L/min) of patients in HFNC/LTOT and LTOT groups at rest were 1.53 (0.95) and 1.64 (1.00), respectively (*p* = 0.577). The mean (SD) of the total flow rate (L/min) in the patients in HFNC/LTOT was 28.5 (4.57).

### Frequency of COPD exacerbations

The unadjusted sample means (HFNC/LTOT, LTOT) of the frequency of a) moderate and severe, b) all-severity, and c) severe-only COPD exacerbations were (1.0, 2.5), (3.8, 5.3), and (0.3, 0.5), respectively. Figure E1 shows histograms of the frequencies of a), b), and c).

Three GLRM models were used to evaluate the frequencies of a), b), and c)-type COPD exacerbations for the primary endpoint, as well as the secondary endpoint 3). The models showed that the adjusted odds ratios (95% CIs) for a), b), and c) were 2.85 (1.48, 5.47), 1.40 (0.91, 2.16), and 1.54 (0.74, 3.22), respectively, when comparing the LTOT and HFNC/LTOT groups at baseline. We found an SSD in the LSM of the frequency of moderate and severe COPD exacerbations between the two treatment groups, with a *p*-value of 0.002. The *p*-values for treatment effects for b) and c) were 0.126 and 0.250, respectively.

### Time to first moderate/severe COPD exacerbation and OS

Figure 1 shows Kaplan-Meier curves for the time to first moderate/severe COPD exacerbation and the OS. A log-rank test for the time to first moderate/severe COPD exacerbation rejected the null hypothesis that the two survival curves were identical (*p* = 0.032). The median survival time (95% CI) for the LTOT group was 25 weeks (14.1, 47.4); however, the HFNC/LTOT group did not reach the median survival time. In contrast, we could not reject the null hypothesis for OS (*p* = 0.947), and both treatment groups did not reach the median survival time. Two patients in each treatment group died during this study.

**Figure 1.**
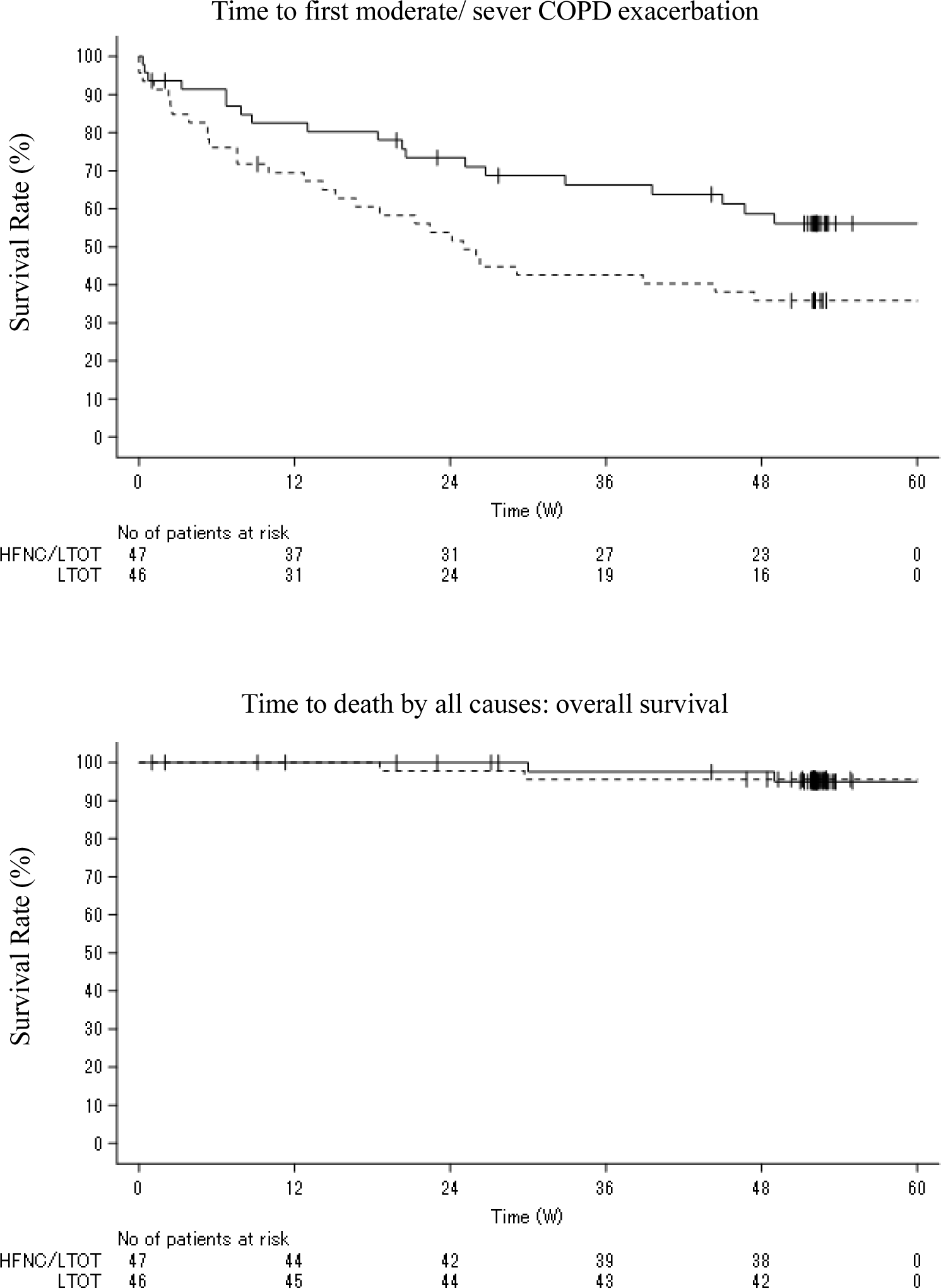
Time to first moderate/severe COPD exacerbation and overall survival (OS). The upper and lower graphs show Kaplan-Meier curves for time to first moderate/severe COPD exacerbation and OS, respectively. The solid and broken lines indicate the HFNC/LTOT and LTOT-alone groups, respectively.

### SGRQ-C (total, symptom, activity, and impact scores)

LSMs with standard errors for total, symptom, activity, and impact scores using the SGRQ-C scale at 12, 24, and 52 weeks are shown in Figure 2. There were no SSDs in LSMs for any scores at any observation time points between the two treatment groups, except for in the impact score at 12 weeks (*p* = 0.028). The LSMs (95% CIs) of the total impact scores for the HFNC/LTOT and LTOT groups at this time point were -3.57 (-8.19, 1.04) and 3.81 (-0.75, 8.36), respectively,

**Figure 2.**
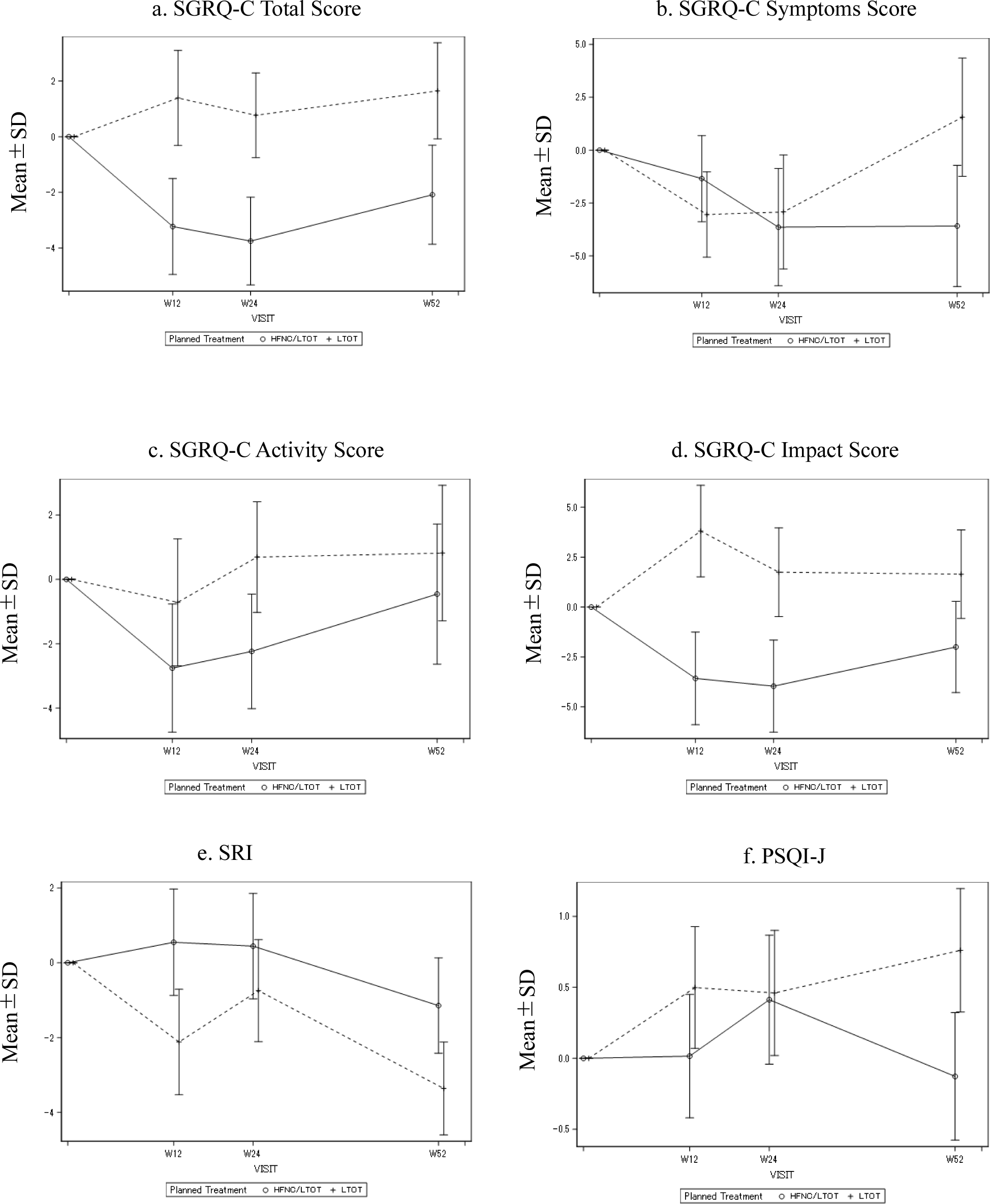
Least-squared means (LSMs) with standard errors of health-related quality of life (HRQOL) scores. Top-left: St. George’s Respiratory Questionnaire (SGRQ-C) total score, top-right: SGRQ-C symptom score, middle-left: SGRQ-C activity score, bottom-left: Severe Respiratory Insufficiency Scale (SRI) score, and bottom-right: Pittsburgh Sleep Quality Index (PSQI-J) score. Solid and broken lines indicate the high-flow nasal cannula oxygen therapy (HFNC)/long-term oxygen therapy (LTOT) and LTOT-alone groups, respectively.

### QALYs

Using ANCOVA, we compared the QALYs between treatment groups, adjusting for the baseline EQ-5D-5L score and treatments; however, there were no SSDs in QALYs between the treatment groups (*p* = 0.270).

### HRQOL

For SRI and PSQI-J scores, we compared the LSMs between the treatment groups at 12, 24, and 52 weeks (Figure 2). We did not find any SSDs in the LSMs of changes in SRI or PSQI-J scores at any observation time points between the two groups. The *p*-values for SRI scores at 12, 24, and 52 weeks were 0.187, 0.547, and 0.218, respectively. Similarly, the *p*-values for PSQI-J scores at 12, 24, and 52 weeks were 0.432, 0.940, and 0.159, respectively.

The Fisher’s exact test was used to examine differences between mMRC categorical scale patterns between the treatment groups at 12, 24, and 52 weeks. There were no SSDs at baseline, 12, 24, or 52 weeks, with *p*-values of *p* = 0.365, *p* = 0.775, *p* = 0.852, and *p* = 0.922, respectively.

### Arterial blood gas analyses

Figure 3 and Table E2 show box plots and descriptive statistics for arterial blood gas measures, respectively. The *t*-test was used to compare sample means (unadjusted means) of the changes in arterial blood gas measurements between the treatment groups at 12, 24, and 52 weeks. There were no SSDs in the mean values, except for in the PaCO2 at 12 weeks (*p* = 0.039); however, we could not confirm this SSD using the LSM with the MMRM technique (*p* = 0.058).

**Figure 3.**
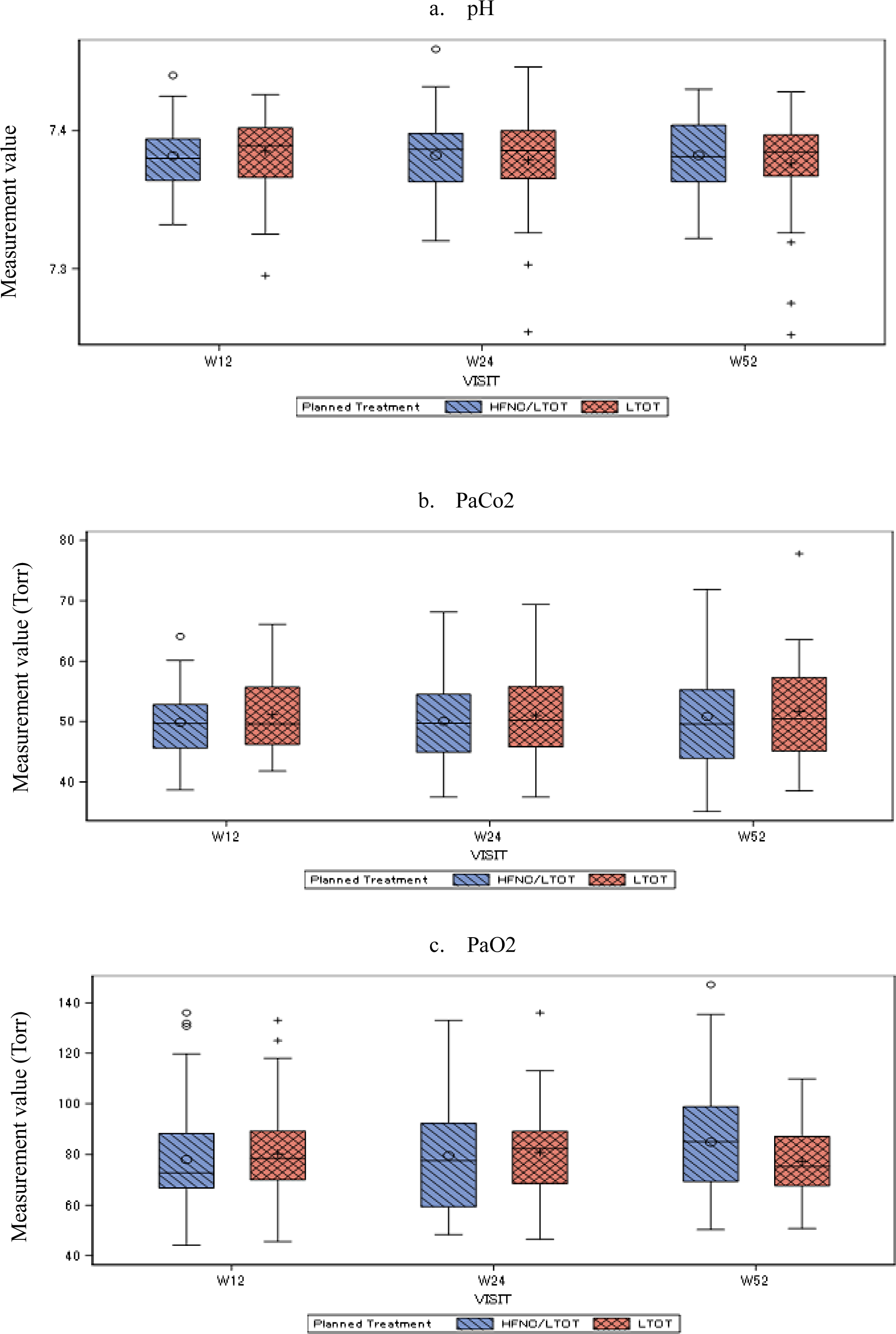
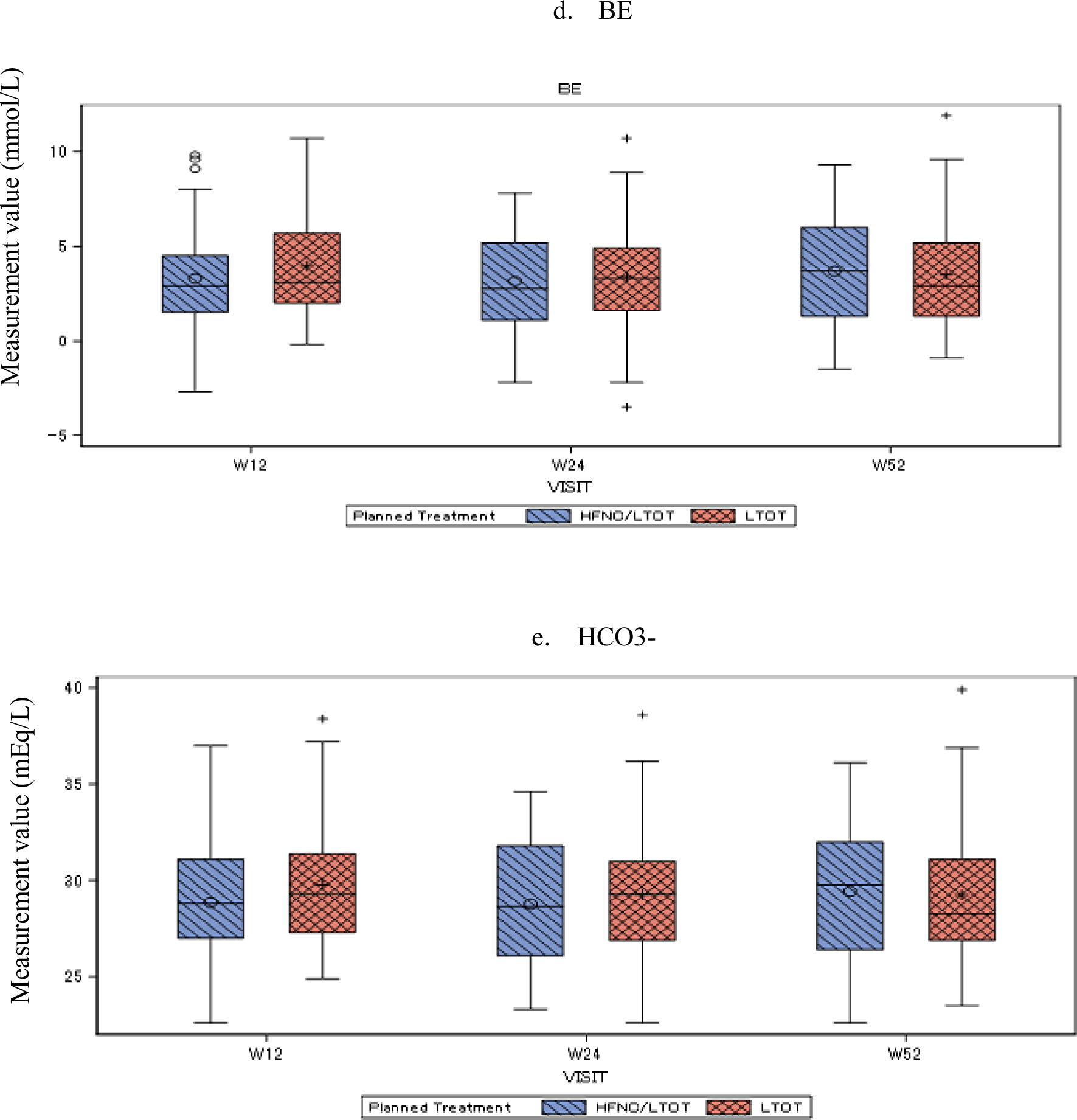
Box plots for arterial blood gas analyses. The left and right boxes summarize the unadjusted observational data at each time point in the high-flow nasal cannula oxygen therapy (HFNC)/long-term oxygen-therapy (LTOT) and LTOT-alone groups, respectively.

### SpO2

At baseline, the sample means ± standard errors of SpO2 for the HFNC/LTOT and LTOT groups were 94.8 ± 3.15 and 95.3 ± 2.51, respectively, without a SSD (*p* = 0.331).

Figure 4 shows the LSM changes in SpO2 at observation time points from 4 to 52 weeks. We repeated the measurement 14 times but did not find any SSDs between the treatment groups at any observation time point except for at 52 weeks (*p* = 0.010). The LSMs ± standard errors of the changes in SpO2 in the HFNC/LTOT and LTOT groups at that week were 1.01 ± 0.33% and -0.20 ± 0.32%, respectively.

**Figure 4.**
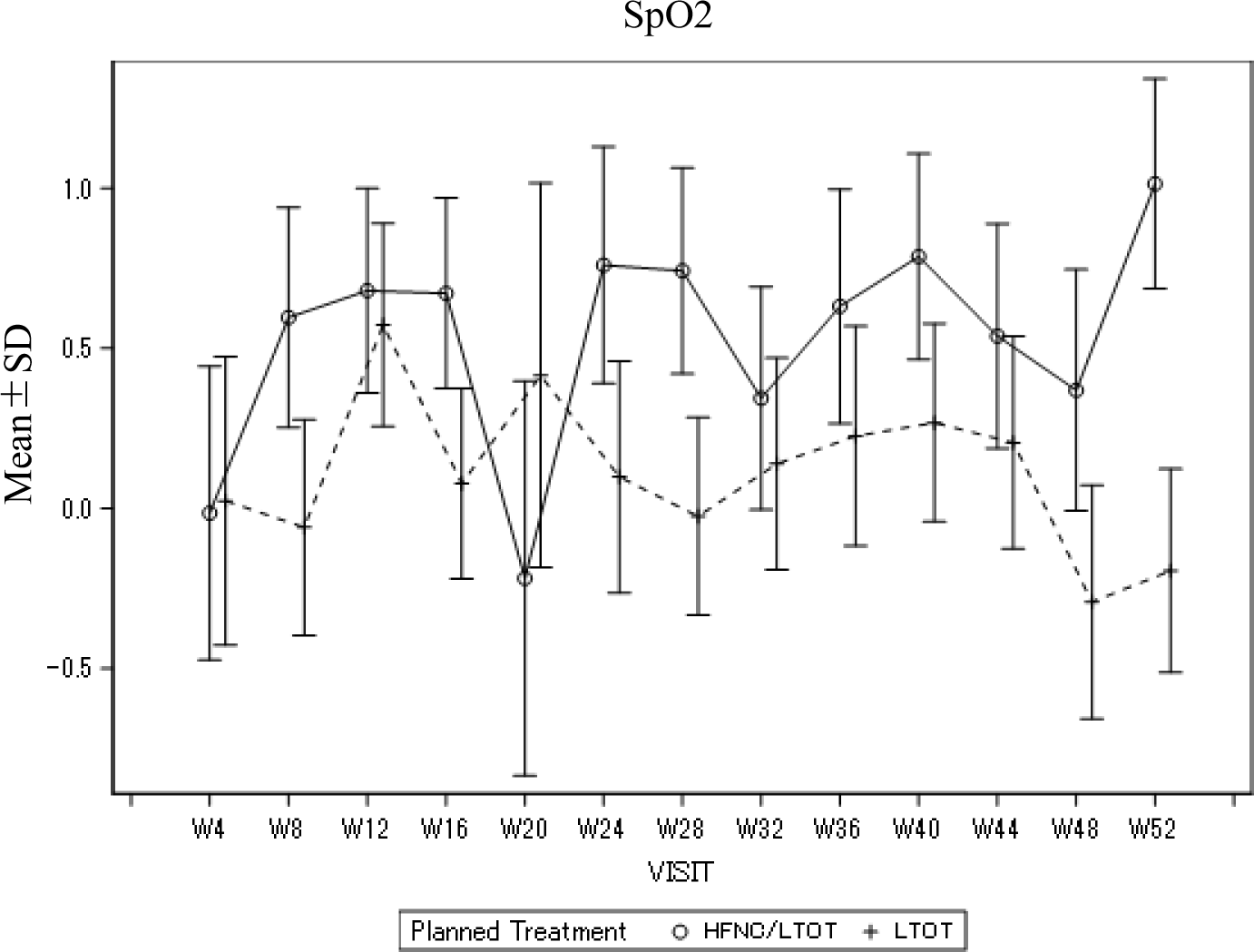
Least-squared means (LSM) with standard errors for peripheral oxygen saturation (SpO2). The solid and broken lines indicate the high-flow nasal cannula oxygen therapy (HFNC)/long-term oxygen therapy (LTOT) and LTOT-alone groups, respectively.

### Pulmonary function

Table 2 shows descriptive statistics at baseline and at 12, 24, and 52 weeks, with *p*-values obtained using the *t*-test to compare the sample means of changes between the treatment groups. Although there were no SSDs at baseline, we found SSDs in the FVC and %FVC at 24 weeks and in the FEV1 and %FEV1 at 12 weeks (*p* = 0.0173, *p* = 0.0148, *p* = 0.0448, *p* = 0.0263, respectively). The mean ± standard error of these items in the HFNC/LTOT and LTOT groups were 2.14 ± 0.54 L, 2.07 ± 0.62 L, 66.74 ± 15.74%, 65.41 ± 17.79%, 0.68 ± 0.23 L, 0.65 ± 0.21 L, 26.89 ± 9.23%, and 26.86 ± 9.32%, respectively. There were no other SSDs between the treatment groups at any observation time.

**Table 2.** Pulmonary function at baseline and at 12, 24, and 52 weeks *P*-values show the results of *t*-tests used to compare the sample means of changes in parameters between treatment groups. The changes are the differences between baseline values and the observed values under treatments. HFNC, high-flow nasal cannula oxygen therapy; LTOT, long-term oxygen therapy; VC, vital capacity; FVC, forced vital capacity; FEV, forced expiratory volume in 1 second; DLCO, diffusing capacity for carbon monoxide; W, week; SD, standard deviation

| Category | Group |  | Baseline | W12 | W24 | W52 |
| --- | --- | --- | --- | --- | --- | --- |
| VC | HFNC/LTOT | Mean (SD) | 2.19 (0.51) | 2.31 (0.54) | 2.28 (0.50) | 2.24 (0.50) |
|  |  | N | 46 | 41 | 38 | 36 |
|  | LTOT | Mean (SD) | 2.25 (0.54) | 2.25 (0.60) | 2.32 (0.62) | 2.27 (0.61) |
|  |  | N | 43 | 42 | 40 | 39 |
|  | *P*-value |  |  | 0.853 | 0.898 | 0.351 |
| %VC | HFNC/LTOT | Mean (SD) | 67.25 (14.31) | 70.27 (13.89) | 69.56 (14.33) | 68.2 (14.38) |
|  |  | N | 46 | 41 | 38 | 36 |
|  | LTOT | Mean (SD) | 69.73 (14.68) | 70.21 (17.72) | 71.66 (17.47) | 70.6 (17.37) |
|  |  | N | 43 | 42 | 40 | 39 |
|  | *P*-value |  |  | 0.742 | 0.783 | 0.420 |
| FVC | HFNC/LTOT | Mean (SD) | 2.01 (0.54) | 2.14 (0.56) | 2.14 (0.54) | 2.05 (0.56) |
|  |  | N | 46 | 41 | 38 | 36 |
|  | LTOT | Mean (SD) | 2.12 (0.62) | 2.05 (0.63) | 2.07 (0.62) | 2.07 (0.63) |
|  |  | N | 45 | 42 | 40 | 39 |
|  | *P*-value |  |  | 0.154 | 0.017 | 0.888 |
| %FVC | HFNC/LTOT | Mean (SD) | 63.12 (15.80) | 67.01 (15.83) | 66.74 (15.74) | 64.02 (16.39) |
|  |  | N | 46 | 41 | 38 | 36 |
|  | LTOT | Mean (SD) | 66.89 (17.09) | 65.55 (18.57) | 65.41 (17.79) | 65.92 (18.69) |
|  |  | N | 45 | 42 | 40 | 39 |
|  | *P*-value |  |  | 0.127 | 0.015 | 0.917 |
| FEV1 | HFNC/LTOT | Mean (SD) | 0.64 (0.22) | 0.68 (0.23) | 0.67 (0.26) | 0.66 (0.25) |
|  |  | N | 46 | 41 | 38 | 36 |
|  | LTOT | Mean (SD) | 0.66 (0.19) | 0.65 (0.21) | 0.68 (0.24) | 0.65 (0.21) |
|  |  | N | 46 | 42 | 40 | 39 |
|  | *P*-value |  |  | 0.045 | 0.606 | 0.265 |
| %FEV1 | HFNC/LTOT | Mean (SD) | 25.59 (8.40) | 26.89 (9.23) | 26.41 (10.07) | 26.27 (9.62) |
|  |  | N | 46 | 41 | 38 | 36 |
|  | LTOT | Mean (SD) | 27.08 (8.94) | 26.86 (9.32) | 27.31 (8.98) | 26.69 (9.42) |
|  |  | N | 46 | 42 | 40 | 39 |
|  | *P*-value |  |  | 0.026 | 0.462 | 0.388 |
| FEV1/FVC | HFNC/LTOT | Mean (SD) | 32.65 (8.86) | 32.5 (8.48) | 31.53 (8.53) | 32.76 (8.77) |
|  |  | N | 46 | 41 | 38 | 36 |
|  | LTOT | Mean (SD) | 32.53 (10.14) | 32.43 (9.63) | 34.13 (14.03) | 32.14 (10.52) |
|  |  | N | 45 | 42 | 40 | 39 |
|  | *P*-value |  |  | 0.181 | 0.349 | 0.202 |

**Table 2 Continued**
| Category | Group |  | Baseline | W12 | W24 | W52 |
| --- | --- | --- | --- | --- | --- | --- |
| DLCO | HFNC/LTOT | Mean (SD) | 6.9 (2.42) | 6.78 (2.42) | 7 (2.69) | 6.9 (2.21) |
|  |  | N | 33 | 36 | 31 | 30 |
|  | LTOT | Mean (SD) | 6.74 (3.27) | 6.75 (3.79) | 7.07 (3.69) | 6.18 (3.32) |
|  |  | N | 24 | 28 | 29 | 28 |
|  | *P*-value |  |  | 0.415 | 0.763 | 0.850 |
| %DLCO | HFNC/LTOT | Mean (SD) | 52.27 (22.32) | 49.31 (24.08) | 54.26 (26.38) | 49.27 (18.73) |
|  |  | N | 33 | 36 | 31 | 29 |
|  | LTOT | Mean (SD) | 47.91 (20.51) | 49.09 (26.17) | 50.81 (23.67) | 44.36 (22.98) |
|  |  | N | 24 | 28 | 29 | 28 |
|  | *P*-value |  |  | 0.286 | 0.695 | 0.918 |

### 6MWT

At 12, 24, and 52 weeks, there was no SDDs between the treatment groups for the mean values of changes in walk distance (*p* = 0.059, 0.678, 0.177, respectively), changes of SpO2 between pre-test and post-test (*p* = 0.609, 0.434, 0.937, respectively), or the modified Borg scale (*p* = 0.427, 0.845, 0.306, respectively).

The mean (SD) of the differences in the walk distance between baseline and 12, 24, 52 weeks in the HFNC/LTOT group was 12.48 (44.26), 14.01 (62.83), and 8.8 (72.17), respectively; in LTOT groups, -11.13 (60.18), 8.19 (52.94), and -12.85 (55.34), respectively. Thus, the *p*-values to test the mean between treatments at 12, 24, 52 weeks were 0.059, 0.678, 0.177, respectively.

### Time to noninvasive positive-pressure ventilation (NPPV)

Only three patients in each treatment group needed long-term NPPV, and the means ± standard errors of the duration of NPPV use for the HFNC/LTOT and LTOT groups were 188.0± 8.9 days and 234.7 ± 178.3 days, respectively. A univariate test was unable to be performed due to the small number of patients with this event.

### Adverse events

The majority of AEs of at least a moderate degree appeared in several patients in both treatment groups. The types of AEs that had more than a 5% frequency in the HFNC/LTOT and LTOT groups were infections and infestations (26.5% and 32.0%, respectively) and respiratory, thoracic, or mediastinal disorders (38.8% and 42.0%, respectively) (Table E3).

## DISCUSSION

In this study, we evaluated the efficacy and safety of HFNC in addition to LTOT alone in hypercapnic COPD patients over 52 weeks, and found that HFNC/LTOT could reduce the COPD exacerbation frequency and prolong the duration between moderate or severe COPD exacerbations. There were several variables with SSDs between the treatment groups, including the SGRQ-C impact score (in the 12^th^ week), SpO2 (in the 52^nd^ week), FVC and %FVC (in the 24^th^ week), and FEV1 and %FEV1 (in the 12^th^ week). Although there were no SSDs in arterial blood gas parameters, HRQOL scores, QALYs, or 6MWT results, there were favoring trends for HFNC in all those parameters. AEs were infrequent in both groups, suggesting that HFNC/LTOT is a safe treatment. To our knowledge, this is the first study to demonstrate that HFNC can be effective and safe for the most severe COPD patients with chronic hypercapnic respiratory failure.

Previous studies conducted in similar settings support our finding that domiciliary HFNC can reduce the frequency of moderate/severe COPD exacerbations. In a randomized trial, Storgaard et al. reported that 12 months of HFNC in a home setting could reduce the frequencies of COPD exacerbations and hospitalizations in chronic hypoxic COPD patients, with hypercapnia in only half of patients.[19] In another randomized trial, Rea et al. demonstrated that HFNC significantly reduced the number of exacerbation days and increased the time to first exacerbation in a mixed population of COPD and bronchiectasis patients.[32]

The current study, however, differs in several important ways from these previous ones. First, our study involved the novel rationale that the most severely sick subgroup of COPD patients would be the most likely to benefit from domiciliary HFNC. Therefore, the current study enrolled only severe COPD patients with chronic hypercapnia and hypoxia who were using LTOT at baseline, which is an important difference from the previous trials. Second, adherence to HFNC was relatively good in the current trial (mean [SD] of 7.3 [3.0] hours/day) compared with that in previous trials. HFNC was used only 1–2 hours/day in Rea’s study and was continued during the observation period in only half of the patients in Storgaard’s study. Lastly, the number of institutions was much larger in the current study, which enabled our results to be generalizable to a larger population.

Despite the reduction in the number of moderate/severe COPD exacerbations in this study, there were only modest effects on physiological parameters and HRQOL. Although there were favorable changes in the SGRQ-C impact score, SpO2, and some pulmonary function parameters, they were only transitory. Therefore, we cannot clearly explain the observed improvement in COPD exacerbations based on these changes. The improvement in the SpO2 at 52 weeks was remarkable; however, a longer observation time is necessary to confirm if this improvement is long lasting. Nevertheless, it may be possible to assume that the observed reduction in COPD exacerbations resulted from the combined effects of those favorable physiological changes.

Although worsening airflow limitation (a lower FEV1) is associated with an increased risk of COPD exacerbations, many other potential contributors have also been reported, including a prior history of exacerbations, lower quality of life, chronic hypercapnia, impaired sleep quality, and chronic mucous hypersecretion.[33–36] Considering those predisposing factors, it is reasonable to associate the combination of multiple factors (although the contribution of each one is small) with the observed reduction in the number of COPD exacerbations. In addition, it may be possible that our method of assessing COPD exacerbations using a daily diary was more sensitive than other parameters for detecting changes in disease. Moreover, the improvement of PFTs found in the HFNC group might have resulted from the reduction of COPD exacerbation, which could accelerate lung function decline.[37] In contrast, several parameters, such as HRQOL and the 6MWT, might take a longer period of time to improve, because they have been established over a long period of time.

In spite of the significant improvements found in SGRQ-C and PaCO2 in our pilot trial,[38] there were numerical improvements of only 1–2 mmHg in the PaCO2 and by only 4–8 points in the SGRQ-C total score, without significant differences, in this study. These improvements were smaller than those reported in previous studies, in which HFNC was demonstrated to improve the PaCO2 by 3–8 mmHg[39–42] and the SGRQ-C total score by 7.8 points.[38] The lack of a significant and continuous improvement in these parameters in the present study is thought to have occurred because we evaluated participants for a longer period of time. In addition, this study included only the most severe COPD patients; therefore, the results were influenced not only by the treatments but also by the progression of COPD. We also evaluated blood gas measurements in the daytime several hours after cessation of HFNC and did not evaluate carbon dioxide during night. These factors may have led to an underestimation of improvements in physiological parameters.

The mechanisms by which HFNC leads to long-term improvement are unclear and remain to be elucidated; however, HFNC is believed to have two major advantages over conventional oxygen delivery systems, resulting in better physiological effects. First, HFNC can effectively provide humidified and heated gas to the airways, leading to enhanced lung mucociliary clearance.[13] Mucociliary clearance has been well established as a first-line defense mechanism of the bronchial tree.[43] The most common cause of a COPD exacerbation is an infection of the lungs or airways; thus, it is reasonable to assume that HFNC can reduce the number of COPD exacerbations by influencing the retention of airway secretions. Moreover, improvements in mucociliary clearance can enhance recruitment and reduce patient respiratory workload.

Another beneficial physiological effect of HFNC is its flushing of the anatomical dead space with the assistance of a positive airway pressure effect, resulting in a lower workload when breathing.[10–13] Many studies have shown that HFNC improved breathing patterns and reduced PaCO2 in stable COPD patients.[38, 41, 44] The combination of these mechanisms may contribute to the beneficial outcomes observed in this trial. Moreover, HFNC can improve swallowing dysfunction[45] which may cause aspiration pneumonia or exacerbations.[46]

The total flow rate of HFNC is of real concern in clinical settings. In an interventional study, flow rate-dependent improvements were observed in the breathing patterns and PaCO2 of COPD patients when changing from 20 L/min up to 50 L/min of a total flow rate.[39] In the short term, a higher flow rate has not been shown to decrease patient comfort;[47] however, in many long-term trials of COPD patients, HFNC was administered mainly at 20–30 L/min,[19, 38, 42] taking into account the balance between effect and comfort. In this trial, the total flow rate was initially targeted for a relatively high flow rate of 30–40 L/min. The observed flow rate, however, was around 30 L/min, which might be due to patients’ comfort levels.

This study had several limitations. First, the lack of a double-blind design was an issue. Unfortunately, the use of a sham device was not possible because it was difficult to blind patients to flow, heat, and humidity, with both patients and clinicians being able to identify a sham device. Instead, the presence of a COPD exacerbation, which was the primary outcome, was diagnosed based on participants’ diaries by a central review panel who were blinded to treatment allocation. Second, we did not adjust the significance level for repeated measurements because our study had an explanatory purpose. However, if we consider the Bonferroni correction for significance levels, the levels should be 0.0125 and 0.004 for the SGRQ-C impact score and pulmonary function testing and the SpO2, respectively. Accordingly, these significance levels were less than all the *p*-values observed in repeated measurements.

## CONCLUSION

In stable hypercapnic patients with a recent history of COPD exacerbations, HFNC reduced the frequency of COPD exacerbations and prolonged the duration between moderate or severe COPD exacerbations over the 52-week study period. However, we could not clearly explain the mechanisms underlying this improvement using our data.

## Data Availability

Data are available upon reasonable request.

## ACKNOWLEDGEMENTS

Kazuma Nagata is the guarantor of the content of the manuscript, including the data and analysis. All the authors contributed substantially to all phases of the study.

The study was sponsored by the Foundation for Biomedical Research and Innovation at Kobe, with funding from Teijin Pharma Limited under the study contract. The funding source of this study participated in writing the study protocol but had no role in data collection, data management, data analysis, or data interpretation. Kikuchi T, a medical statistician, and Nakagawa Y, a programmer, both of whom analyzed study data, are employees of the Foundation for Biomedical Research and Innovation at Kobe. However, the funding source did not review the analyses nor the current manuscript, and all the work was independent from the funder.

English language editing service was provided by Editage (www.editage.jp).

## Table and Figure Legends

**Figure E1.**
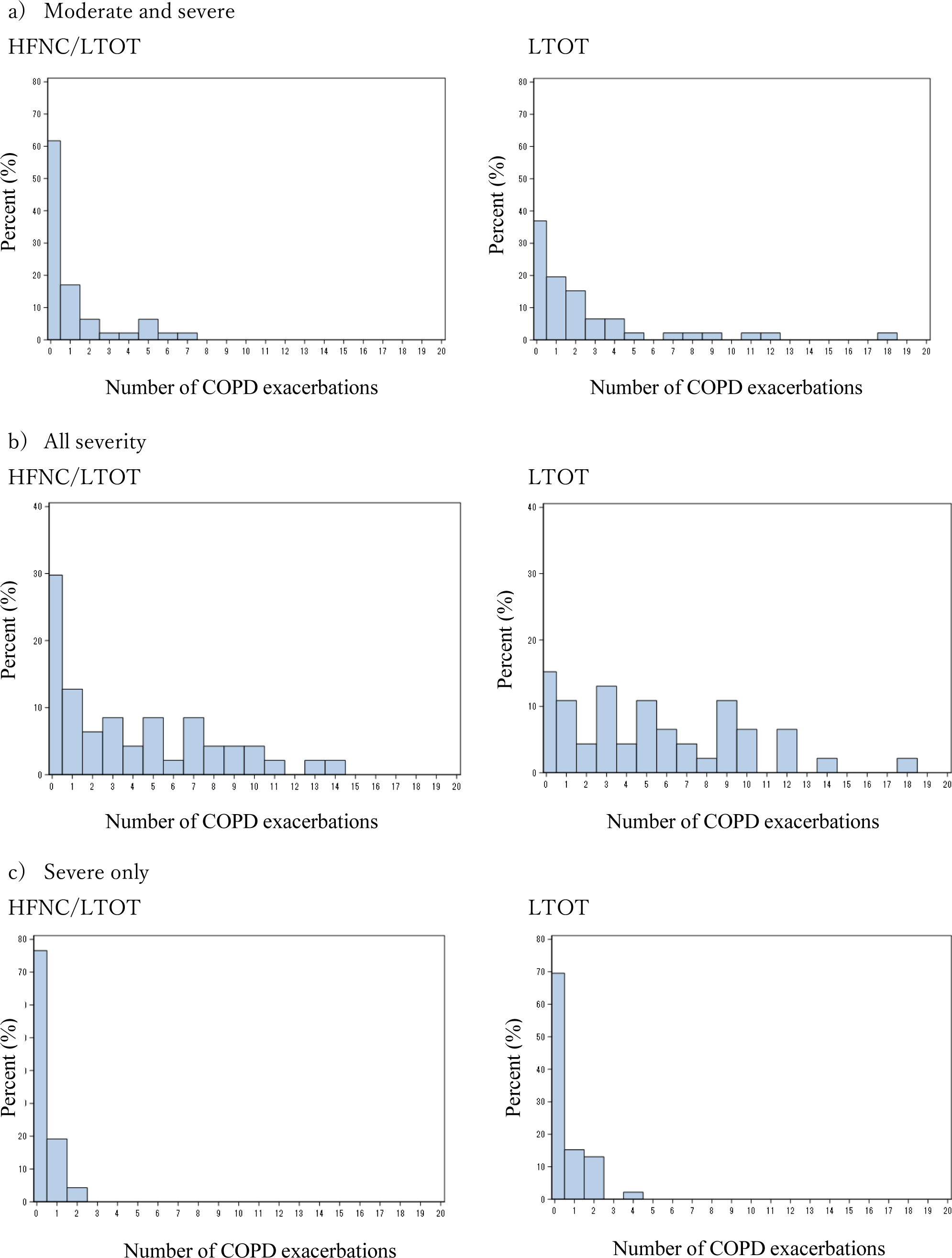
Histograms of chronic obstructive pulmonary disease (COPD) exacerbation frequency. The top, middle, and bottom figures show the frequency of “moderate and severe,” “all-severity,” and “severe-only” COPD exacerbations, respectively. The left and right columns represent the high-flow nasal cannula oxygen therapy (HFNC)/long-term oxygen therapy (LTOT) and LTOT-alone groups, respectively.

**Table E1.** Inclusion and exclusion criteria

|  |
| --- |
| *Inclusion criteria* |
| (1) Men and women aged 40 years or older |
| (2) Diagnosed with chronic obstructive pulmonary disease (COPD) |
| (3) Global Initiative for Obstructive Lung Disease [20] stages 2–4 |
| (4) Receiving long-term oxygen therapy for at least 16 hours per day for at least 1 month prior to providing informed consent |
| (5) Hypercapnic respiratory failure ($\text{PaCO}_2 \geq 45$ mmHg and $\text{pH} \geq 7.35$) at the time of screening |
| (6) Having a COPD exacerbation (moderate or severe; judged by the investigators) within the past 1 year |
| (7) Willing to provide written informed consent to participate in this study |
| *Exclusion criteria* |
| (1) Having severe and unstable comorbidities or an active malignancy |
| (2) History of or highly suspected to have obstructive sleep apnea syndrome |
| (3) Diagnosis of asthma |
| (4) Having a COPD exacerbation within the previous 4 weeks |
| (5) Nocturnal noninvasive ventilation use within the prior 4 weeks or high-flow nasal cannula oxygen therapy within the past 1 year |
| (6) Cognitive impairment or a psychiatric disorder |
| (7) Considered ineligible by the investigator |

**Table E2.** Arterial blood gas analyses This table includes descriptive statistics for arterial blood gas analyses at baseline and at 12, 24, and 52 weeks. *P*-values show the results of *t*-tests used to compare sample means of changes in parameters between treatment groups. The changes represent the difference between baseline values and the observed values under treatments. HFNC, high-flow nasal cannula oxygen therapy; LTOT, long-term oxygen therapy; W, week; SD, standard deviation

| Category | Group |  | Baseline | W12 | W24 | W52 |
| --- | --- | --- | --- | --- | --- | --- |
| pH | HFNC/LTOT | Mean | 7.38 | 7.38 | 7.38 | 7.38 |
|  |  | SD | 0.02 | 0.02 | 0.03 | 0.03 |
|  |  | N | 47 | 41 | 40 | 37 |
|  | LTOT | Mean | 7.39 | 7.39 | 7.38 | 7.38 |
|  |  | SD | 0.02 | 0.03 | 0.03 | 0.04 |
|  |  | N | 46 | 43 | 42 | 38 |
|  | *P*-value |  | . | 0.3838 | 0.1844 | 0.1178 |
| PaCO<sub>2</sub> | HFNC/LTOT | Mean | 51.38 | 49.87 | 50.04 | 50.87 |
|  |  | SD | 4.96 | 6.05 | 6.7 | 8.28 |
|  |  | N | 47 | 41 | 40 | 37 |
|  | LTOT | Mean | 50.50 | 51.09 | 51.00 | 51.65 |
|  |  | SD | 5.03 | 6.37 | 6.85 | 8.57 |
|  |  | N | 46 | 43 | 42 | 38 |
|  | *P*-value |  | . | 0.0386 | 0.1813 | 0.5202 |
| PaO<sub>2</sub> | HFNC/LTOT | Mean | 80.37 | 77.96 | 79.52 | 84.82 |
|  |  | SD | 21.75 | 21.84 | 22.2 | 23.36 |
|  |  | N | 47 | 41 | 40 | 37 |
|  | LTOT | Mean | 84.10 | 80.35 | 80.80 | 77.37 |
|  |  | SD | 21.85 | 18.79 | 17.96 | 14.53 |
|  |  | N | 46 | 43 | 42 | 38 |
|  | *P*-value |  | . | 0.5828 | 0.5213 | 0.0625 |
| HCO<sub>3</sub><sup>-</sup> | HFNC/LTOT | Mean | 29.74 | 28.88 | 28.77 | 29.44 |
|  |  | SD | 2.67 | 3.21 | 3.06 | 3.65 |
|  |  | N | 47 | 41 | 40 | 37 |
|  | LTOT | Mean | 29.72 | 29.79 | 29.27 | 29.24 |
|  |  | SD | 2.74 | 3.34 | 3.33 | 3.67 |
|  |  | N | 46 | 43 | 42 | 38 |
|  | *P*-value |  | . | 0.1245 | 0.5044 | 0.53 |
| BE | HFNC/LTOT | Mean | 3.95 | 3.30 | 3.16 | 3.69 |
|  |  | SD | 2.34 | 2.86 | 2.52 | 2.87 |
|  |  | N | 47 | 41 | 40 | 37 |
|  | LTOT | Mean | 3.90 | 3.95 | 3.39 | 3.51 |
|  |  | SD | 2.37 | 2.74 | 2.75 | 3.02 |
|  |  | N | 45 | 43 | 42 | 38 |
|  | *P*-value |  | . | 0.1709 | 0.6579 | 0.5405 |

**Table E3.** Adverse events This table shows the frequency of adverse events of at least a moderate degree. HFNC, high-flow nasal cannula oxygen therapy; LTOT, long-term oxygen therapy; SOC, system organ class; PT, preferred term; SAE, severe adverse event; CI, confidence interval

| SOC Name | PT Name | HFNC/LTOT |  |  | LTOT |  |  |
| --- | --- | --- | --- | --- | --- | --- | --- |
|  |  | n (%) | 95% CI | SAE | n (%) | 95% CI | SAE |
| Cardiac disorders |  | 3 (6.1) | 1.660–11.394 | 2 | 2 (4.0) | 1.660–11.394 | 1 |
|  | Cardiac failure congestive | 1 (2.0) | 0.052–10.854 |  | 0 (0.0) | - |  |
|  | Cardio-respiratory arrest | 1 (2.0) | 0.052–10.854 | 1 | 0 (0.0) | - |  |
|  | Palpitations | 1 (2.0) | 0.052–10.854 | 1 | 0 (0.0) | - |  |
|  | Tachycardia | 0 (0.0) | - |  | 2 (4.0) | 0.488–13.714 | 1 |
| Ear and labyrinth disorders |  | 1 (2.0) | 0.052–10.854 |  | 0 (0.0) | - |  |
|  | Positional vertigo | 1 (2.0) | 0.052–10.854 |  | 0 (0.0) | - |  |
| Gastrointestinal disorders |  | 0 (0.0) | - |  | 1 (2.0) | 0.051–10.647 |  |
|  | Large intestinal polyp | 0 (0.0) | - |  | 1 (2.0) | 0.051–10.647 |  |
| General disorders and administration site conditions |  | 2 (4.1) | 0.498–13.979 |  | 0 (0.0) | - |  |
|  | Pyrexia | 1 (2.0) | 0.052–10.854 |  | 0 (0.0) | - |  |
|  | Performance status decrease | 1 (2.0) | 0.052–10.854 |  | 0 (0.0) | - |  |
| Hepatobiliary disorders |  | 1 (2.0) | 0.052–10.854 |  | 0 (0.0) | - |  |
|  | Cholecystitis | 1 (2.0) | 0.052–10.854 |  | 0 (0.0) | - |  |
|  | Cholecystitis acute | 1 (2.0) | 0.052–10.854 |  | 0 (0.0) | - |  |
| Infections and infestations |  | 13 (26.5) | 20.574–39.291 | 5 | 16 (32.0) | 20.574–39.291 | 5 |
|  | Bronchopulmonary aspergillosis | 1 (2.0) | 0.052–10.854 |  | 0 (0.0) | - |  |
|  | Influenza | 1 (2.0) | 0.629–8.601 |  | 2 (4.0) | 0.629–8.601 | 1 |
|  | Nasopharyngitis | 2 (4.1) | 0.629–8.601 |  | 1 (2.0) | 0.629–8.601 | 1 |
|  | Pneumonia | 9 (18.4) | 14.481–31.689 | 4 | 13 (26.0) | 14.481–31.689 | 4 |
|  | Septic shock | 1 (2.0) | 0.052–10.854 | 1 | 0 (0.0) | - |  |
|  | Urinary tract infection | 2 (4.1) | 0.629–8.601 | 1 | 1 (2.0) | 0.629–8.601 |  |
|  | Respiratory tract infection | 1 (2.0) | 0.052–10.854 | 1 | 0 (0.0) | - |  |

Table E3. Continued
| SOC Name | PT Name | HFNC/LTOT |  |  | LTOT |  |  |
| --- | --- | --- | --- | --- | --- | --- | --- |
|  |  | n (%) | 95% CI | SAE | n (%) | 95% CI | SAE |
| Metabolism and nutrition disorders |  | 0 (0.0) | - |  | 1 (2.0) | 0.051–10.647 |  |
|  | Dehydration | 0 (0.0) | - |  | 1 (2.0) | 0.051–10.647 |  |
| Musculoskeletal and connective tissue disorders |  | 2 (4.1) | 1.112–10.023 |  | 2 (4.0) | 1.112–10.023 |  |
|  | Arthralgia | 1 (2.0) | 0.246–7.108 |  | 1 (2.0) | 0.246–7.108 |  |
|  | Tendonitis | 0 (0.0) | - |  | 1 (2.0) | 0.051–10.647 |  |
|  | Immobilization syndrome | 1 (2.0) | 0.052–10.854 |  | 0 (0.0) | - |  |
| Nervous system disorders |  | 0 (0.0) | - |  | 1 (2.0) | 0.051–10.647 |  |
|  | Cognitive disorder | 0 (0.0) | - |  | 1 (2.0) | 0.051–10.647 |  |
| Renal and urinary disorders |  | 2 (4.1) | 0.498–13.979 | 1 | 0 (0.0) | - |  |
|  | Pollakiuria | 1 (2.0) | 0.052–10.854 |  | 0 (0.0) | - |  |
|  | Renal failure | 1 (2.0) | 0.052–10.854 | 1 | 0 (0.0) | - |  |
| Respiratory, thoracic, and mediastinal disorders |  | 19 (38.8) | 30.657–50.741 | 11 | 21 (42.0) | 30.657–50.741 | 10 |
|  | Chronic obstructive pulmonary disease | 15 (30.6) | 24.176–43.518 | 9 | 18 (36.0) | 24.176–43.518 | 8 |
|  | Dyspnea | 1 (2.0) | 0.052–10.854 |  | 0 (0.0) | - |  |
|  | Hemoptysis | 1 (2.0) | 0.052–10.854 | 1 | 0 (0.0) | - |  |
|  | Pneumothorax | 2 (4.1) | 1.660–11.394 | 2 | 3 (6.0) | 1.660–11.394 | 2 |
|  | Respiratory failure | 1 (2.0) | 0.052–10.854 | 1 | 0 (0.0) | - |  |
|  | Upper respiratory tract inflammation | 1 (2.0) | 0.246–7.108 |  | 1 (2.0) | 0.246–7.108 |  |
|  | Lung cyst | 1 (2.0) | 0.052–10.854 | 1 | 0 (0.0) | - |  |
| Skin and subcutaneous tissue disorders |  | 0 (0.0) | - |  | 1 (2.0) | 0.051–10.647 |  |
|  | Urticaria | 0 (0.0) | - |  | 1 (2.0) | 0.051–10.647 |  |

Table E3. Continued
| SOC Name | PT Name | HFNC/LTOT |  |  | LTOT |  |  |
| --- | --- | --- | --- | --- | --- | --- | --- |
|  |  | n (%) | 95% CI | SAE | n (%) | 95% CI | SAE |
| Vascular disorders |  | 1 (2.0) | 0.052–10.854 |  | 0 (0.0) | - |  |
|  | Hypertension | 1 (2.0) | 0.052–10.854 |  | 0 (0.0) | - |  |

## Supplemental Appendix

### Study Organization

#### Principal Investigator

Keisuke Tomii, Department of Respiratory Medicine, Kobe City Medical Center General Hospital, Kobe, Japan

#### Project Director

Kazuma Nagata, Department of Respiratory Medicine, Kobe City Medical Center General Hospital, Kobe, Japan.

#### Steering Committee

Keisuke Tomii, Department of Respiratory Medicine, Kobe City Medical Center General Hospital, Kobe, Japan.

Kazuma Nagata, Department of Respiratory Medicine, Kobe City Medical Center General Hospital, Kobe, Japan.

Fumiaki Tokioka, Department of Respiratory Medicine, Kurashiki Central Hospital, Kurashiki, Japan Joe Shindo, Department of Respiratory Medicine, Ogaki Municipal Hospital, Gifu, Japan.

Akira Shiraki, Department of Respiratory Medicine, Ogaki Municipal Hospital, Gifu, Japan. Motonari Fukui, Respiratory Disease Center, Tazuke Kofukai Foundation, Medical Research Institute, Kitano Hospital, Osaka, Japan.

Takamasa Kitajima, Respiratory Disease Center, Tazuke Kofukai Foundation, Medical Research Institute, Kitano Hospital, Osaka, Japan.

Naohiko Chohnabayashi, Division of Pulmonary Medicine, Thoracic Center, St. Luke’s International Hospital, Tokyo, Japan.

Torahiko Jinta, Division of Pulmonary Medicine, Thoracic Center, St. Luke’s International Hospital, Tokyo, Japan.

Ryosuke Tsugitomi, Division of Pulmonary Medicine, Thoracic Center, St. Luke’s International Hospital, Tokyo, Japan.

Takeo Horie, Department of Respiratory Medicine, Japanese Red Cross Maebashi Hospital, Maebashi, Japan.

Toru Kadowaki, Department of Pulmonary Medicine, National Hospital Organization Matsue Medical Center, Matsue, Japan.

Akira Watanabe, Department of Respiratory Medicine, National Hospital Organization Ehime Medical Center, Ehime, Japan.

Susumu Sato, Department of Respiratory Medicine, Graduate School of Medicine, Kyoto University, Kyoto, Japan.

#### Trial Statistician

Takashi Kikuchi, Department of Medical Statistics, Translational Research Center for Medical Innovation, Foundation for Biomedical Research and Innovation at Kobe, Kobe, Japan.

#### Data Center

Translational Research Center for Medical Innovation, Foundation for Biomedical Research and Innovation at Kobe, Kobe, Japan.

#### Efficacy and Safety Evaluation Committee

Yoshihiro Nishimura, Division of Respiratory Medicine, Department of Internal Medicine, Kobe University Graduate School of Medicine, Kobe, Japan.

Hisashi Onishi, Department of Respiratory Medicine, Akashi Medical Center, Akashi, Japan. Kazuhiro Kajimoto, Department of Respiratory Medicine, Kobe Red Cross Hospital, Kobe, Japan.

#### FLOCOP study investigators

Osamu Hataji (Matsusaka Municipal Hospital), Akira Watanabe (National Hospital Organization Ehime Medical Center), Michiko Tsuchiya (Rakuwakai Otowa Hospital), Miyuki Okuda (Federation of National Public Service Personnel Mutual Aid Association Hirakata Kohsai Hospital), Satoshi Fuke (KKR Sapporo Medical Center), Takeo Horie (Japanese Red Cross Maebashi Hospital), Tomomasa Tsuboi (National Hospital Organization Minami Kyoto Hospital), Toru Kadowaki (National Hospital Organization Matsue Medical Center), Sayaka Tachibana (Ehime Prefectural Central Hospital), Shohei Takata (National Hospital Organization Fukuokahigashi Medical Center), Tadashi Ishida (Kurashiki Central Hospital), Michiko Kagajo (Ogaki Municipal Hospital), Hisayuki Osoreda (NHO Yamaguchi-Ube Medical Center), Motonari Fukui (Kitano Hospital Tazuke Kofukai Medical Research Institute), Takashi Nishimura (Kyoto Katsura Hospital), Iwao Gohma (Sakai City Medical Center), Nobuhito Kishimoto (Takamastu Municipal Hospital), Yoshihiro Mori (KKR Takamatsu Hospital), Yoshikazu Inoue, Toru Arai, Takayuki Takimoto (Kinki-Chuo Chest Medical Center), Hideo Kita (Takatsuki Red-Cross Hospital), Makoto Yoshida (National Hospital Organization Fukuoka National Hospital), Takao Kamimori (Yodogawa Christian Hospital), Kenichi Minami (Ishikiriseiki Hospital), Takashi Niwa (Kanagawa Prefectural Hospital Organization Kanagawa Cardiovascular and Respiratory Center), Torahiko Jinta (St.Luke’s International Hospital), Kenichi Takahashi (Kishiwada City Hospital), Tomoo Kishaba (Okinawa Chubu Hospital), Yasuo Kohashi (HARUHI Respiratory Medical Hospital), Masayuki Nakayama (Jichi Medical University Hospital), Toru Tsuda (Kirigaoka Tsuda Hospital), Jiro Terada (Chiba University Hospital), Shinyu Izumi (National Center for Global Health and Medicine), Hiroki Nishine (St. Marianna University School of Medicine), Hiroyuki Nakamura (Sakaide City Hospital), Hiroyasu Kobayashi (Suzuka General Hospital), Yasuhiro Kondoh (Tosei General Hospital), Ichiro Nakachi (Saiseikai Utsunomiya Hospital), Takashi Ogasawara (Hamamatsu Medical Center), Hiroaki Momi, Koichi Takagi, Hiromasa Inoue (Kagoshima University Hospital), Teruaki Nishiuma (Kakogawa Central City Hospital), Yasuto Nakatsumi (Kanazawa Municipal Hospital)

## REFERENCES

1. Viegi, G., et al., Definition, epidemiology and natural history of COPD. Eur Respir J, 2007. 30(5): p. 993–1013.

2. Foucher, P., et al., Relative survival analysis of 252 patients with COPD receiving long-term oxygen therapy. Chest, 1998. 113(6): p. 1580–1587.

3. García-Rivero, J.L., et al., Risk Factors of Poor Outcomes after Admission for a COPD Exacerbation: Multivariate Logistic Predictive Models. Copd, 2017. 14(2): p. 164–169.

4. Kohnlein, T., et al., Non-invasive positive pressure ventilation for the treatment of severe stable chronic obstructive pulmonary disease: a prospective, multicentre, randomised, controlled clinical trial. Lancet Respir Med, 2014. 2(9): p. 698–705.

5. Dreher, M., et al., High-intensity versus low-intensity non-invasive ventilation in patients with stable hypercapnic COPD: a randomised crossover trial. Thorax, 2010. 65(4): p. 303–308.

6. Murphy, P.B., et al., Effect of Home Noninvasive Ventilation With Oxygen Therapy vs Oxygen Therapy Alone on Hospital Readmission or Death After an Acute COPD Exacerbation: A Randomized Clinical Trial. Jama, 2017. 317(21): p. 2177–2186.

7. Cheng, S.L., V.L. Chan, and C.M. Chu, Compliance with home non-invasive ventilation. Respirology, 2012. 17(4): p. 735–736.

8. Nava, S., P. Navalesi, and C. Gregoretti, Interfaces and humidification for noninvasive mechanical ventilation. Respir Care, 2009. 54(1): p. 71–84.

9. Fanfulla, F., et al., Effect of sleep on patient/ventilator asynchrony in patients undergoing chronic non-invasive mechanical ventilation. Respir Med, 2007. 101(8): p. 1702–1707.

10. Dysart, K., et al., Research in high flow therapy: mechanisms of action. Respir Med, 2009. 103(10): p. 1400–1405.

11. Gotera, C., et al., Clinical evidence on high flow oxygen therapy and active humidification in adults. Rev Port Pneumol, 2013. 19(5): p. 217–227.

12. Ricard, J.D., High flow nasal oxygen in acute respiratory failure. Minerva Anestesiol, 2012. 78(7): p. 836–841.

13. Hasani, A., et al., Domiciliary humidification improves lung mucociliary clearance in patients with bronchiectasis. Chron Respir Dis, 2008. 5(2): p. 81–86.

14. Frat, J.P., et al., High-flow oxygen through nasal cannula in acute hypoxemic respiratory failure. N Engl J Med, 2015. 372(23): p. 2185–2196.

15. Jones, P.G., et al., Randomized Controlled Trial of Humidified High-Flow Nasal Oxygen for Acute Respiratory Distress in the Emergency Department: The HOT-ER Study. Respir Care, 2016. 61(3): p. 291–299.

16. Hernández, G., et al., Effect of Postextubation High-Flow Nasal Cannula vs Conventional Oxygen Therapy on Reintubation in Low-Risk Patients: A Randomized Clinical Trial. Jama, 2016. 315(13): p. 1354–1361.

17. Fernandez, R., et al., High-flow nasal cannula to prevent postextubation respiratory failure in high-risk non-hypercapnic patients: a randomized multicenter trial. Ann Intensive Care, 2017. 7(1): p. 47.

18. Bonnevie, T., et al., Nasal High Flow for Stable Patients with Chronic Obstructive Pulmonary Disease: A Systematic Review and Meta-Analysis. Copd, 2019. 16(5-6): p. 368–377.

19. Storgaard, L.H., et al., Long-term effects of oxygen-enriched high-flow nasal cannula treatment in COPD patients with chronic hypoxemic respiratory failure. Int J Chron Obstruct Pulmon Dis, 2018. 13: p. 1195–1205.

20. Vogelmeier, C.F., et al., Global Strategy for the Diagnosis, Management, and Prevention of Chronic Obstructive Lung Disease 2017 Report. GOLD Executive Summary. Am J Respir Crit Care Med, 2017. 195(5): p. 557–582.

21. Terada, K., et al., Impact of gastro-oesophageal reflux disease symptoms on COPD exacerbation. Thorax, 2008. 63(11): p. 951–955.

22. Anthonisen, N.R., et al., Antibiotic therapy in exacerbations of chronic obstructive pulmonary disease. Ann Intern Med, 1987. 106(2): p. 196–204.

23. Meguro, M., et al., Development and Validation of an Improved, COPD-Specific Version of the St. George Respiratory Questionnaire. Chest, 2007. 132(2): p. 456–463.

24. Windisch, W., et al., The Severe Respiratory Insufficiency (SRI) Questionnaire: a specific measure of health-related quality of life in patients receiving home mechanical ventilation. J Clin Epidemiol, 2003. 56(8): p. 752–759.

25. Herdman, M., et al., Development and preliminary testing of the new five-level version of EQ-5D (EQ-5D-5L). Qual Life Res, 2011. 20(10): p. 1727–1736.

26. Doi, Y., et al., Psychometric assessment of subjective sleep quality using the Japanese version of the Pittsburgh Sleep Quality Index (PSQI-J) in psychiatric disordered and control subjects. Psychiatry Res, 2000. 97(2-3): p. 165–172.

27. Buysse, D.J., et al., The Pittsburgh Sleep Quality Index: a new instrument for psychiatric practice and research. Psychiatry Res, 1989. 28(2): p. 193–213.

28. Bestall, J.C., et al., Usefulness of the Medical Research Council (MRC) dyspnoea scale as a measure of disability in patients with chronic obstructive pulmonary disease. Thorax, 1999. 54(7): p. 581–586.

29. Miller, M.R., et al., Standardisation of spirometry. Eur Respir J, 2005. 26(2): p. 319–338.

30. Society, T.J.R., Reference values of spirogram and arterial blood gas levels in Japanese. Ann Jpn Respir Soc, 2001. 39: p. 1–17.

31. Laboratories, A.C.o.P.S.f.C.P.F., ATS statement: guidelines for the six-minute walk test. Am J Respir Crit Care Med, 2002. 166(1): p. 111–117.

32. Rea, H., et al., The clinical utility of long-term humidification therapy in chronic airway disease. Respir Med, 2010. 104(4): p. 525–533.

33. Suzuki, M., et al., Clinical features and determinants of COPD exacerbation in the Hokkaido COPD cohort study. Eur Respir J, 2014. 43(5): p. 1289–1297.

34. Rothnie, K.J., et al., Natural History of Chronic Obstructive Pulmonary Disease Exacerbations in a General Practice-based Population with Chronic Obstructive Pulmonary Disease. Am J Respir Crit Care Med, 2018. 198(4): p. 464–471.

35. Chu, C.M., et al., Readmission rates and life threatening events in COPD survivors treated with non-invasive ventilation for acute hypercapnic respiratory failure. Thorax, 2004. 59(12): p. 1020–1025.

36. Shorofsky, M., et al., Impaired Sleep Quality in COPD Is Associated With Exacerbations: The CanCOLD Cohort Study. Chest, 2019. 156(5): p. 852–863.

37. Halpin, D.M.G., et al., Effect of a single exacerbation on decline in lung function in COPD. Respir Med, 2017. 128: p. 85–91.

38. Nagata, K., et al., Domiciliary High-Flow Nasal Cannula Oxygen Therapy for Patients with Stable Hypercapnic Chronic Obstructive Pulmonary Disease. A Multicenter Randomized Crossover Trial. Ann Am Thorac Soc, 2018. 15(4): p. 432–439.

39. Braunlich, J., M. Kohler, and H. Wirtz, Nasal highflow improves ventilation in patients with COPD. Int J Chron Obstruct Pulmon Dis, 2016. 11: p. 1077–1085.

40. Braunlich, J., et al., Effects of nasal high flow on ventilation in volunteers, COPD and idiopathic pulmonary fibrosis patients. Respiration, 2013. 85(4): p. 319–325.

41. Fraser, J.F., et al., Nasal high flow oxygen therapy in patients with COPD reduces respiratory rate and tissue carbon dioxide while increasing tidal and end-expiratory lung volumes: a randomised crossover trial. Thorax, 2016. 71(8): p. 759–761.

42. Braunlich, J., H.J. Seyfarth, and H. Wirtz, Nasal High-flow versus non-invasive ventilation in stable hypercapnic COPD: a preliminary report. Multidiscip Respir Med, 2015. 10(1): p. 27.

43. Wanner, A., M. Salathe, and T.G. O’Riordan, Mucociliary clearance in the airways. Am J Respir Crit Care Med, 1996. 154(6 Pt 1): p. 1868-1902.

44. McKinstry, S., et al., Nasal high flow therapy and PtCO2 in stable COPD: A randomized controlled cross-over trial. Respirology, 2018. 23(4): p. 378–384.

45. Sanuki, T., et al., Effect of nasal high-flow oxygen therapy on the swallowing reflex: an in vivo volunteer study. Clin Oral Investig, 2017. 21(3): p. 915–920.

46. Nagami, S., et al., Breathing-swallowing discoordination is associated with frequent exacerbations of COPD. BMJ Open Respir Res, 2017. 4(1): p. e000202.

47. Mauri, T., et al., Impact of flow and temperature on patient comfort during respiratory support by high-flow nasal cannula. Crit Care, 2018. 22(1): p. 120.

